# Genetic score associations with birthweight in preterm-born infants compared with term-born infants

**DOI:** 10.1101/2025.01.21.25320880

**Authors:** Robin N Beaumont, Sarah J Kotecha, Shannon J Simpson, Sailesh Kotecha, Rachel M Freathy

## Abstract

**Objective:** In preterm infants, lower birthweight correlates with a higher risk of neonatal complications. Understanding the factors influencing birthweight in these infants is important as it may guide future antenatal and perinatal care. Genetic variants account for at least one-quarter of variation in birthweight in term-born infants, but the genetic contribution to birthweight in preterm infants is not well understood. We aimed to compare genetic score associations with birthweight in a well-powered sample of preterm infants with those in term-born infants.

**Study design:** We used linear regression to test the association between birth weight and fetal genetic scores for birthweight (BW-GS) in a total of 1,416 preterm, singleton and 15,253 term, singleton infants. Analyses, adjusted for ancestry principal components were performed within each of 4 datasets and meta-analysed.

**Results:** In term-born infants: a 1 SD higher BW-GS was associated with a 1.20 (95% CI 1.10-1.29) SD higher birthweight. In preterm infants, there was also strong evidence of association, but with a smaller effect size (0.76SD (0.41-1.11) higher birthweight per 1-SD higher BW-GS). In preterms, when stratifying by gestational duration, we found that the associations strengthened with increasing gestational duration.

**Conclusions:** Genetic scores composed of variants identified in term-born infants also influenced birthweight in preterm infants. However, the associations had smaller effects in preterms and were weaker at earlier gestations. This suggests that while many of the same genetic factors influence birthweight in preterm and term-born infants, other factors (environmental, placental, different genetic) may be more important in preterms. Future well-powered studies are required to investigate this.

## Introduction

Infants born below the 3^rd^ percentile of weight for gestational age at term (37 or more weeks of gestation) are at higher risks of illness and death than those in the middle quartiles, but in preterm births (before 37 weeks), those risks are proportional to the reduction in birthweight percentile, rather than being limited to the most severely undergrown infants^1^. Fetal growth restriction is both an important cause of iatrogenic preterm birth and an independent risk factor for spontaneous preterm birth^2^. A clear understanding of the factors influencing birthweight in preterm-born infants is therefore important for future prediction and prevention of neonatal complications.

Fetal and maternal genetic factors are important contributors to birthweight. Estimates of heritability based on genome-wide association studies suggest that 39.8% of birthweight variation is attributable to common genetic variation, with contributions from the fetal genome (28.5%), the maternal genome (7.6%) and the covariance between them (3.7%)^3^. However, to date, genetic associations with birthweight have been investigated mostly in term-born infants, with a very limited number focused on preterm births.

Studies that have examined genetic associations with birthweight or estimated fetal weight before term have noted similarities with studies of term birthweight. For example, the common birthweight-associated variant with the largest effect identified from the first fetal genome-wide association study (GWAS) of birthweight in term-born infants was similarly associated with birthweight in a study of 1,194 preterm-born infants^4^. Another study assessing fetal growth using ultrasound scans showed that a fetal genetic score of 59 birthweight-associated variants, identified from GWAS consisting mostly of term-born infants, was associated with estimated fetal weight at 30 weeks of gestation, but not at 20 weeks in 5,374 pregnancies^5^.

In this study, we aimed to compare the associations between birthweight and fetal genetic scores for birthweight, length or ponderal index, in a well-powered sample of 1,416 singleton, preterm-born infants, with those of 15,253 infants born at term.

## Methods

### Cohort Descriptions

#### Preterm-specific Cohorts (RHiNO, WALHIP and PICSI)

The respiratory health outcomes in neonates (RHiNO) dataset for this analysis consisted of children of European genetic similarity, born at or before 34 weeks of gestation. The Western Australian Lung Health in Prematurity (WALHIP^6^) and the Preterm Inhaled CorticoSteroid Intervention (PICSI^7^) datasets consisted of children and young adults born ≤32 weeks of gestation in Western Australia. Genotyping (total n=669 individuals; 429 RHiNO, 101 WALHIP, 90 PICSI) for these studies was performed together using the Illumina GSA Array v2. Following exclusions (genotype call rate <98%, deviation from Hardy-Weinberg equilibrium (P<1×10-6), and ancestry principal component analysis^8^ outliers), 617 individuals and 559,430 SNPs remained for analysis. Imputation was performed using the Michigan Imputation Server up to the HRC reference panel. Birthweight was available from medical records for 533 individuals with genotype data, and full details of data collection can be found in the RHiNO study protocol^9^ or cohort descriptions^6,7^. Ethical approval was obtained from the local ethics committees.

#### ALSPAC

From the Avon Longitudinal Study of Parents and Children (ALSPAC) a total of 6,943 term (gestational duration >=37 weeks) and 328 preterm offspring with genotype and birthweight data were available for analysis. The study protocol, genotyping and imputation have been described previously^10,11^. Ethical approval for the study was obtained from the ALSPAC Ethics and Law Committee and the Local Research Ethics Committees.

#### Millennium Cohort study

The Millennium Cohort study (MCS) is a longitudinal cohort study of children born around the turn of the 21st century, from which 5,854 term- and 440 preterm-born individuals had birthweight and genotype data available for our analysis^12,13^.

#### Born in Bradford

The Born in Bradford (BiB) is a multi-ethnic longitudinal birth study based in Bradford, UK^14^. Data were available for 2,456 and 3229 term-, and 117 and 126 preterm-born participants of European and South Asian genetic similarity, respectively, with both genotype and phenotype data available.

### Phenotype preparation

Birthweights were converted to z-scores using growth charts appropriate to term- or preterm-born infants. For RHiNO, WALHIP, PICSI, MCS, and BiB, the stata (v16) package zanthro was used, with the UKWHO or UKWHO preterm charts^15^. For ALSPAC, birthweight z-scores were calculated adjusting for gestational age and sex using LMSgrowth^16^.

### Construction of genetic scores

Genetic scores (GS) for birthweight (BW), birth length (BL), and ponderal index (PI) were constructed with higher GS corresponding to higher genetically predicted BW, BL or PI respectively. For the BW GS, we used lead SNPs at each of the 190 loci identified by Warrington et al^3^. BL and PI GSs were constructed using the 10 and 7 lead SNPs from the respective analyses by Juliusdottir et al^17^. The BL and PI GS analyses aimed to assess genetic contribution to the skeletal and fat mass components of birthweight, respectively. GSs were calculated using Eq 1 where *w_i_* is the weight, and *g_i_* is the dosage of the effet allele at SNP *i*. GSs for BW were constructed using both raw weights uncorrected for correlation between maternal and fetal genotypes, and using weights from the SEM of Warrington et al, to account for this correlation.

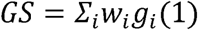

### Association analysis

Associations between each GS and birthweight were tested in term- and preterm-born infants separately using linear regression. Analyses were performed in each cohort separately, adjusting for principal components, and meta-analysed using inverse variance weighted meta-analysis for individuals of European genetic similarity. Associations in South Asian genetic similarity individuals were compared to the European-like meta-analysis results. In both term- and preterm-born infants, we also performed analyses in male and female infants separately. Additionally, we stratified the preterm group into gestational bands of between 34 and 37 weeks, 32 and 34 weeks, and up to 32 weeks, and analyses within each subgroup.

## Results

As expected, we found strong associations between birthweight GS and birthweight in term-born infants: a 1 SD higher BW GS was associated with a 1.20 (95% CI 1.10-1.29) SD higher birthweight (Figure 1). The birthweight genetic score was also associated with birthweight in preterm-born infants. However, the effect size was smaller: a 1 SD higher birthweight GS in the preterm group was associated with a 0.76 (0.41-1.11) SD higher birthweight. Within the preterm group, the association between birthweight GS and birthweight was similar for infants born at 34-36 and 32-33 weeks gestation. However, there was little evidence of association in infants born between 28-31 weeks (Figure 1). Associations were similar between the males and females in both the term and preterm groups (all P>0.05; Figure 1). There was no evidence of heterogeneity between studies in the meta-analysis (all P_bonferroni_>0.05/42).

**Figure 1:**
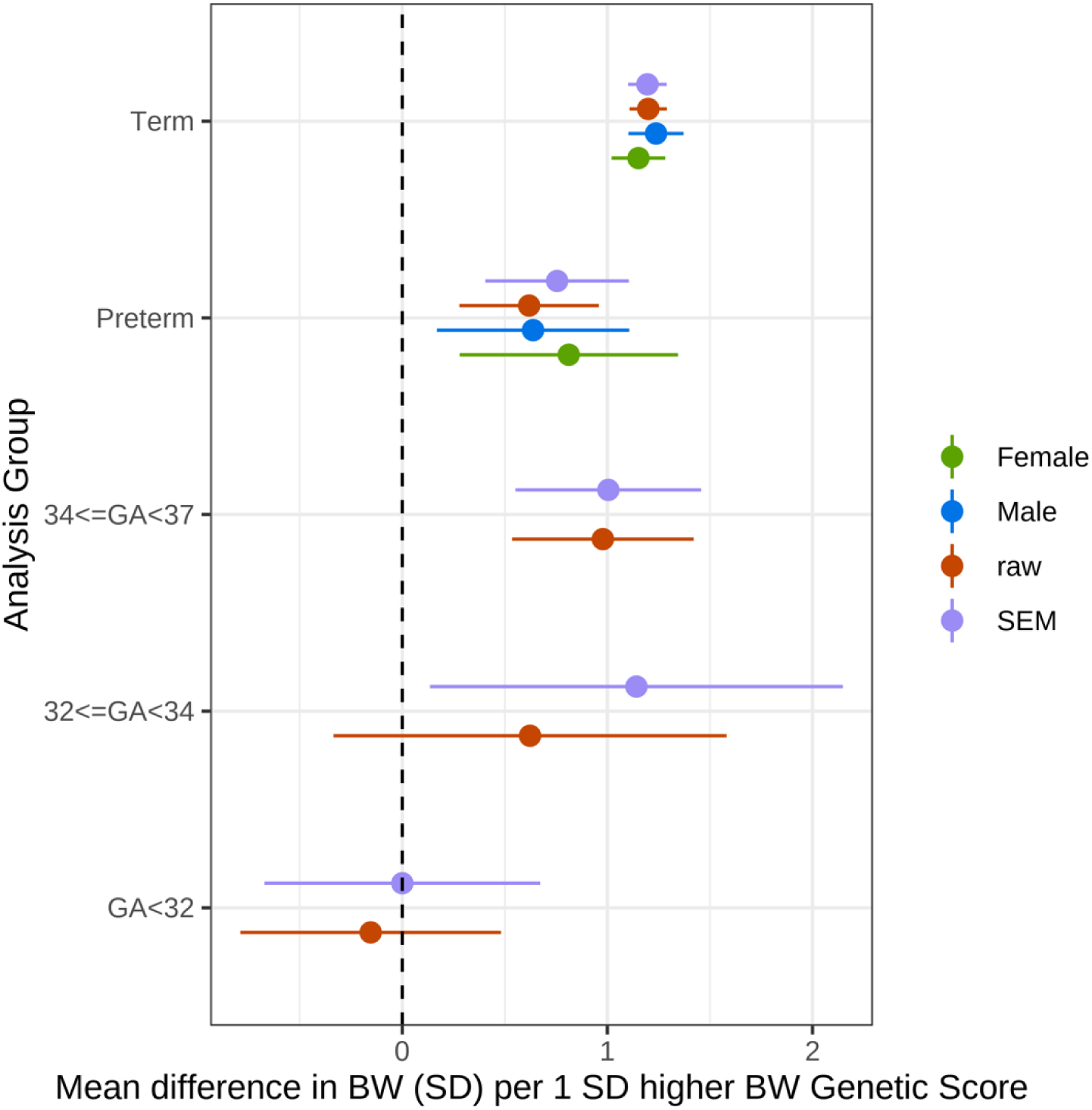
Associations between birthweight GS and birthweight in term- and preterm-born infants. Associations between birthweight GS and birthweight in term- and preterm-born infants from a meta-analysis of 4 samples of European genetic similarity. Total sample sizes: Term n=15,253 (7556 female, 7697 male), Preterm n=1416 (642 female, 774 male), 34<=GA<37 n=755, 32<=GA<34 n=222, GA<32 n=429.

Comparing results in European genetic similarity participants to those of South Asian genetic similarity from BiB, we found that associations beween birthweight GS and birthweight in both term and preterm groups were smaller in South Asian participants (Figure 2). Within the preterm groups, there was no evidence of association between birthweight GS and birthweight, although confidence intervals were wide.

**Figure 2:**
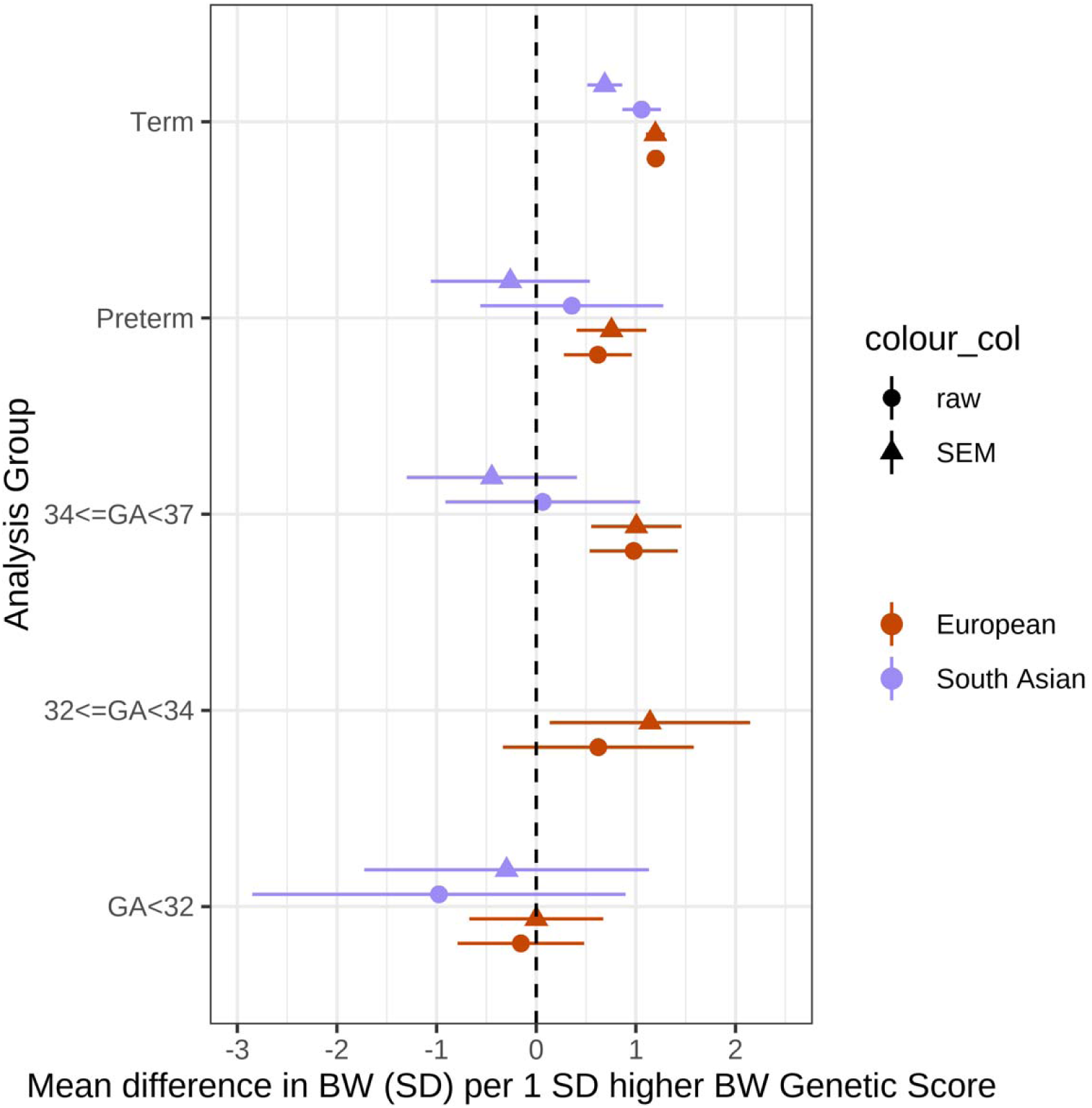
Associations between birthweight GS and birthweight in term- and preterm-born infants in European and South Asian genetic similarity individuals. Associations between birthweight GS and birthweight in term- and preterm-born infants from a meta-analysis of 4 European ancestry studies compared to South Asian genetic similarity individuals from the BiB cohort. Total sample sizes (Pakistani): Term 3229, Preterm 126, 34<=GA<37 107, GA<32 11. There were too few individuals with 32<=GA<34 to perform association analysis.

The ponderal index GS showed strong evidence of association with birthweight in term-born infants (1.04; 95% CI: 0.87-1.22), however, there was little evidence of association in preterm-born infants (0.23; 95% CI: −0.40-0.86; Figure 3). The birth length GS showed strong evidence of association with birthweight at term (0.76; 95% CI: 0.61-0.90; Figure 3), and there was also some evidence for an association of birth length GS in preterm-born infants (0.55; 95% CI: 0.00-1.10). This association was mainly driven by those born late preterm, with strong evidence for an association in infants born at 34-36 weeks gestation (1.31; 95% CI: 0.59-2.04) but little evidence for an association in infants born before 32 weeks (−0.72; 95% CI: −1.77-0.32).

**Figure 3:**
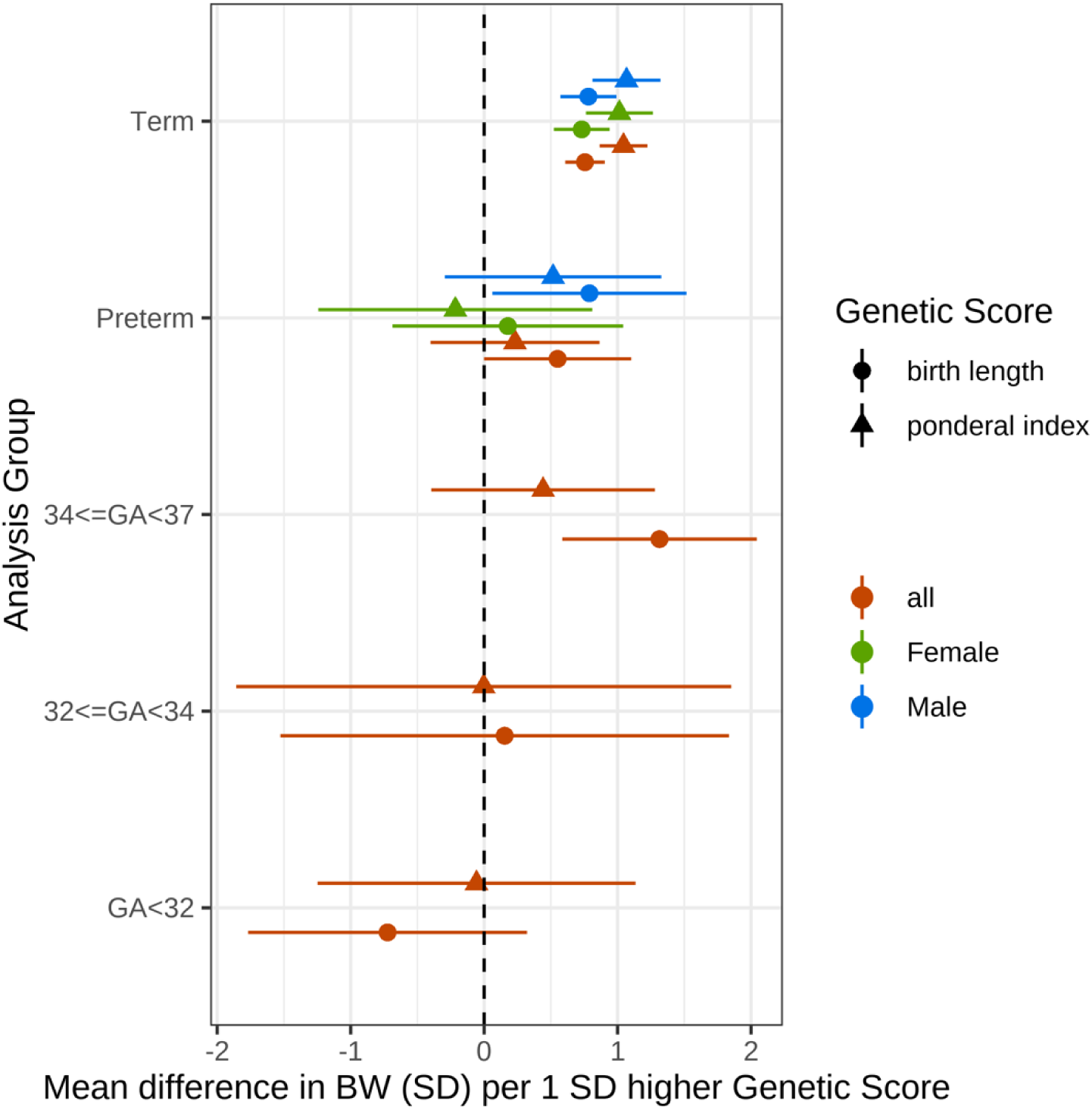
Associations between birth length GS and birthweight in preterm- and term-born infants. Associations between birth length (bl) and ponderal index (pi) GSs and birthweight in term- and preterm-born infants from a meta-analysis of 4 studies. Total sample sizes: Term 15,253 (7556 female, 7697 male), Preterm 1416 (642 female, 774 male), 34<=GA<37 755, 32<=GA<34 222, GA<32 429.

## Discussion

In this study, we found that a genetic score composed of variants each known to have a small effect on birthweight from previous GWAS, was strongly associated with birthweight in both term- and preterm-born infants. The magnitude of this association was smaller in the preterm than in the term sample, and there was little evidence for sex differences in any of the associations.

Our results are similar to those of Vermeulen et al.^5^, who noted that the effect estimate of a birthweight GS association with fetal weight, estimated from ultrasound scan data in singletons, was approximately half that for term birthweight. Our observation that genetic score associations are detectable in both term- and preterm-born infants is also consistent with the similar genetic architecture underlying birthweight in twins, who have shorter gestations and lower birthweights than term-born infants, on average^18^.

The observation that the strength of the association in South Asian individuals was weaker than in European ancestry individuals may reflect the fact that the birthweight GSs, which were discovered in primarily European genetic similarity samples, captured birthweight-associated genetic variation less well in the South Asian individuals due to differing linkage disequilibrium patterns across ancestry groups. It could also reflect the fact that growth standards for both term and preterm babies for which standardized birthweight values were calculated, are more specific to European genetic similarity individuals. These discrepancies highlight the need for better, ancestry-specific growth references to better capture expected growth in these groups, and GSs which are more transferrable to individuals of non-European ancestries.

At earlier gestational ages, we observed attenuation in the magnitude of the genetic score associations with birthweight. Although our stratified sample size was low, limiting statistical power, there was evidence for a difference in effect of the birthweight GS between the 28-31 week and the 34-36 week group (P=0.011). We also found that the ponderal index GS was more weakly associated with birthweight in the preterm vs. the term sample. However, we found no evidence that the associations between the birth length GS and birthweight were different between term- and preterm-born infants. Taken together, these results suggest that the smaller association of birthweight GS with birthweight in preterm infants compared with term-born infants is more likely due to reduced effects on adiposity than to reduced effects on skeletal growth.

Different factors may influence growth at different stages of gestation. For example, fetal insulin-mediated growth occurs mainly during the 3rd trimester as suggested by analyses of infants with absent fetal insulin^19^. Several of the genetic loci making up the birthweight GS are also associated with adult glycemic traits^3^, and our results are consistent with the genetic score effects on fetal insulin-mediated growth becoming more apparent with increasing gestation. It is unlikely that insulin-mediated growth can fully explain this difference, however, because the majority of birthweight-associated genetic loci have not been implicated in glycaemic traits, and the birthweight GS is not associated with cord insulin levels^20^.

The attenuation of the associations at earlier gestational ages suggests that there were more infants in the early preterm groups whose growth differed from their genetic potential. For example, the weaker genetic associations may have been due to restricted fetal growth owing to the impact of environmental factors or poor placental health. In addition, since the variants we used to construct the genetic scores were identified in samples of people almost exclusively born at term^3,17^, it may be that there are some different genetic factors underlying birthweight in preterm-born infants. Additional work focusing on adequately powered preterm-born cohorts will be required to investigate this.

## Acknowledgements

We are extremely grateful to all the participants and their families who took part in all the cohorts, the midwives and research staff for their help in recruiting them, and the whole cohort teams, which included health professionals, interviewers, computer and laboratory technicians, clerical workers, research scientists, volunteers, managers, receptionists and nurses. Born in Bradford is only possible because of the enthusiasm and commitment of the children and parents in BiB. We are grateful to all the participants, health professionals, schools and researchers who have made Born in Bradford happen.

## Data Availability

Please note that the ALSPAC study website contains details of all the data that is available through a fully searchable data dictionary and variable search tool" and reference the following webpage: http://www.bristol.ac.uk/alspac/researchers/our-data/

Scientists are encouraged and able to use BiB data. Data requests are made to the BiB executive using the form available from the study website http://www.borninbradford.nhs.uk (please click on ‘Science and Research’ to access the form). Guidance for researchers and collaborators, the study protocol and the data collection schedule are all available via the website. All requests are carefully considered and accepted where possible.

The MCS data is available from the Centre for Longitudinal Studies Data Access Committee (see here for details: https://cls.ucl.ac.uk/data-access-training/data-access/)

Data are available on reasonable request. All data relevant to the study are included in the article or uploaded as online supplemental information. Data from the RHiNO study are available to research collaborators subject to confidentiality and non-disclosure agreements. Contact Professor SK for any data requests

## Author Contributions

RNB, SJK, SK, RMF contributed to study design; RNB performed the analyses in the study; RNB, SJK, SK, RMF aided in data interpretation and statistical analysis; RNB drafted the original manuscript including all tables and figures. All authors reviewed and edited versions of the manuscript.

## Online Tables

**Table 1:**
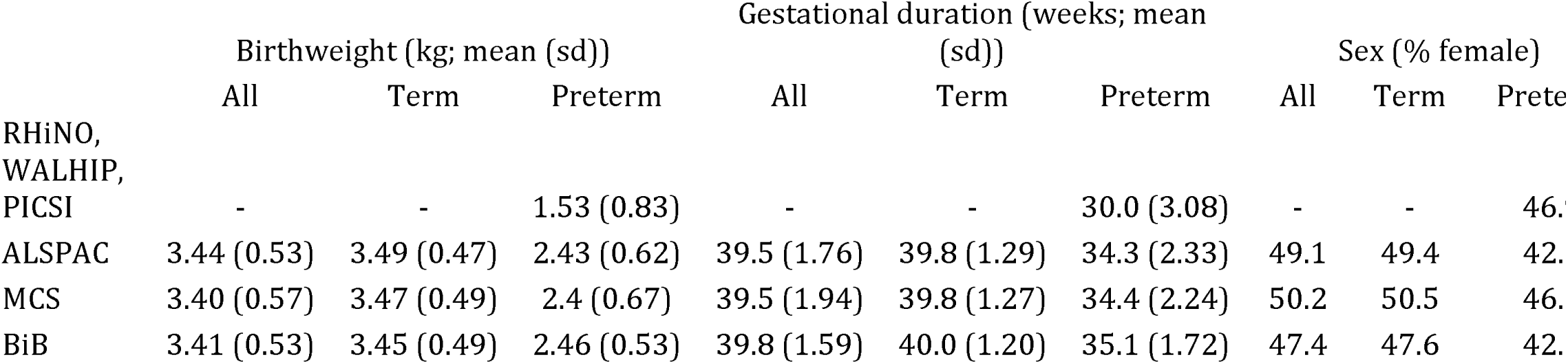
Demographic statistics for included samples.

**Table 2:** Associations between birthweight GS and birthweight in term- and preterm-born infants. For genetic scores, “raw” indicates the score constructed using the marginal effects from the GWAS meta-analysis of the fetal genome vs own BW and “SEM” indicates the estimates from the partitioned fetal effects of the GWAS variants.

| Gestation | Sex | Genetic Score | Beta | SE | P |
| --- | --- | --- | --- | --- | --- |
| 34<=GA<37 | all | raw | 0.9781 | 0.2259 | 1.50E-05 |
| 34<=GA<37 | all | SEM | 1.0047 | 0.231 | 1.36E-05 |
| 32<=GA<34 | all | raw | 0.6229 | 0.4888 | 2.03E-01 |
| 32<=GA<34 | all | SEM | 1.1417 | 0.5134 | 2.62E-02 |
| GA<32 | all | raw | -0.0392 | 0.153 | 7.98E-01 |
| GA<32 | all | SEM | 0.106 | 0.161 | 5.08E-01 |
| Preterm | all | raw | 0.6184 | 0.1736 | 3.67E-04 |
| Preterm | all | SEM | 0.7552 | 0.1786 | 2.35E-05 |
| Preterm | Female | SEM | 0.812 | 0.2717 | 2.81E-03 |
| Preterm | Male | SEM | 0.6378 | 0.2394 | 7.73E-03 |
| Term | all | raw | 1.1993 | 0.0466 | 3.53E-146 |
| Term | all | SEM | 1.1953 | 0.048 | 3.70E-137 |
| Term | Female | SEM | 1.1516 | 0.0669 | 2.50E-66 |
| Term | Male | SEM | 1.2374 | 0.0686 | 9.46E-73 |

**Table 3:** Associations between birth length GS and birthweight in preterm- and term-born infants.

| Genetic Score | Gestation | Beta | SE | P |
| --- | --- | --- | --- | --- |
| ponderal index | 34<=GA<37 | 0.4419 | 0.4274 | 3.01E-01 |
| ponderal index | 32<=GA<34 | -0.0026 | 0.9463 | 9.98E-01 |
| ponderal index | GA<32 | -0.0569 | 0.608 | 9.25E-01 |
| ponderal index | All | 0.9922 | 0.0894 | 1.20E-28 |
| ponderal index | All (Female) | 0.9679 | 0.1264 | 1.91E-14 |
| ponderal index | All (Male) | 1.0038 | 0.1262 | 1.80E-15 |
| ponderal index | Preterm | 0.2314 | 0.323 | 4.74E-01 |
| ponderal index | Preterm (Female) | -0.2156 | 0.5238 | 6.81E-01 |
| ponderal index | Preterm (Male) | 0.5167 | 0.4138 | 2.12E-01 |
| ponderal index | Term | 1.0442 | 0.0913 | 2.57E-30 |
| ponderal index | Term (Female) | 1.0138 | 0.128 | 2.31E-15 |
| ponderal index | Term (Male) | 1.0672 | 0.13 | 2.21E-16 |
| birth length | 34<=GA<37 | 1.3144 | 0.3719 | 4.09E-04 |
| birth length | 32<=GA<34 | 0.154 | 0.8574 | 8.58E-01 |
| birth length | GA<32 | -0.723 | 0.522 | 1.75E-02 |
| birth length | All | 0.7609 | 0.0741 | 9.54E-25 |
| birth length | All (Female) | 0.7367 | 0.1052 | 2.55E-12 |
| birth length | All (Male) | 0.7855 | 0.1041 | 4.51E-14 |
| birth length | Preterm | 0.5507 | 0.2815 | 5.04E-02 |
| birth length | Preterm (Female) | 0.1777 | 0.4409 | 6.87E-01 |
| birth length | Preterm (Male) | 0.7891 | 0.371 | 3.34E-02 |
| birth length | Term | 0.7558 | 0.0756 | 1.53E-23 |
| birth length | Term (Female) | 0.7312 | 0.1066 | 6.92E-12 |
| birth length | Term (Male) | 0.7816 | 0.107 | 2.73E-13 |

## References

1. McIntire, D. D., Bloom, S. L., Casey, B. M. & Leveno, K. J. Birth Weight in Relation to Morbidity and Mortality among Newborn Infants. New England Journal of Medicine 340, 1234–1238 (1999).

2. Melamed, N. et al. FIGO (International Federation of Gynecology and Obstetrics) initiative on fetal growth: Best practice advice for screening, diagnosis, and management of fetal growth restriction. International Journal of Gynecology & Obstetrics 152, 3–57 (2021).

3. Warrington, N. M. et al. Maternal and fetal genetic effects on birth weight and their relevance to cardio-metabolic risk factors. Nat Genet 51, 804–814 (2019).

4. Ryckman, K. K. et al. Replication of a genome-wide association study of birth weight in preterm neonates. J Pediatr 160, 19–24.e4 (2012).

5. Vermeulen, M. J. et al. Influence of genetic variants for birth weight on fetal growth and placental haemodynamics. Arch Dis Child Fetal Neonatal Ed 105, 393–398 (2020).

6. Smith, E. F. et al. Risk factors for poorer respiratory outcomes in adolescents and young adults born preterm. Thorax 78, 1223–1232 (2023).

7. Inhaled corticosteroids to improve lung function in children (aged 6–12 years) who were born very preterm (PICSI): a randomised, double-blind, placebo-controlled trial - The Lancet Child & Adolescent Health. https://www.thelancet.com/journals/lanchi/article/PIIS2352-4642(23)00128-1/abstract.

8. Abraham, G., Qiu, Y. & Inouye, M. FlashPCA2: principal component analysis of Biobank-scale genotype datasets. Bioinformatics 33, 2776–2778 (2017).

9. RHINO: Respiratory Health Outcomes in Neonates. Health Research Authority https://www.hra.nhs.uk/planning-and-improving-research/application-summaries/research-summaries/rhino-respiratory-health-outcomes-in-neonates/.

10. Boyd, A. et al. Cohort Profile: The ‘Children of the 90s’—the index offspring of the Avon Longitudinal Study of Parents and Children. International Journal of Epidemiology 42, 111–127 (2013).

11. Fraser, A., et al. Cohort Profile: The Avon Longitudinal Study of Parents and Children: ALSPAC mothers cohort. International Journal of Epidemiology 42, 97–110 (2013).

12. Connelly, R. & Platt, L. Cohort profile: UK Millennium Cohort Study (MCS). Int J Epidemiol 43, 1719–1725 (2014).

13. University College London, U. I. O. E. Millennium Cohort Study. Preprint at 10.5255/UKDA-SERIES-2000031 (2024).

14. Wright, J. et al. Cohort Profile: The Born in Bradford multi-ethnic family cohort study. International Journal of Epidemiology 42, 978–991 (2013).

15. Cole, T. J., Freeman, J. V. & Preece, M. A. British 1990 growth reference centiles for weight, height, body mass index and head circumference fitted by maximum penalized likelihood. Statistics in Medicine 17, 407–429 (1998).

16. Pan H, Cole T. LMSgrowth version 2.77. 2012. LMSgrowth version 2.77. 2012. www.healthforallchildren.com/shop-base/shop/software/lmsgrowth.

17. Juliusdottir, T. et al. Distinction between the effects of parental and fetal genomes on fetal growth. Nat Genet 53, 1135–1142 (2021).

18. Beck, J. J. et al. Genetic meta-analysis of twin birth weight shows high genetic correlation with singleton birth weight. Human Molecular Genetics 30, 1894–1905 (2021).

19. Hughes, A. E., Franco, E. D., Freathy, R. M., Flanagan, S. E. & Hattersley, A. T. Monogenic disease analysis establishes that fetal insulin accounts for half of human fetal growth. J Clin Invest (2023) doi:10.1172/JCI165402.

20. Hughes, A. E. et al. Fetal Genotype and Maternal Glucose Have Independent and Additive Effects on Birth Weight. Diabetes 67, 1024–1029 (2018).

